# Clinically Meaningful Improvement in the treatment of Negative Symptoms of Schizophrenia: The Case of Roluperidone

**DOI:** 10.64898/2026.09.07.26362403

**Authors:** Jonathan Rabinowitz, Stefan Leucht, Michael Davidson, Remy Luthringer

**Affiliations:** Bar Ilan University, Ramat Gan, Israel; Minerva Neurosciences, Burlington, MA, USA; Technical University of Munich, Munich, Germany; University of Nicosia Medical School, Cyprus

**Keywords:** negative symptoms, schizophrenia, clinical meaningfulness, roluperidone

## Abstract

**Background:** Clinical meaningfulness in schizophrenia trials requires evidence that symptom change is recognizable in clinical practice. Established anchors include a ≥1-point improvement on the CGI-S and a ≥7-to 10-point improvement on the Personal and Social Performance scale (PSP), alongside the ≥20% improvement on the PANSS negative symptom factor score (NSFS) conventionally applied in negative-symptom trials.

**Methods:** These clinically meaningful anchors were applied as benchmarks to pooled 12-week completer data from two randomized, placebo-controlled trials of roluperidone in adults with schizophrenia and moderate to severe negative symptoms.

**Results:** Roluperidone was associated with higher responder rates than placebo on all three endpoints: ≥20% NSFS improvement (66/176 [38%] vs 39/182 [21%]; RR=1.75, 95% CI: 1.25– 2.45; p=0.0008), ≥1-point CGI-S improvement (68/167 [41%] vs 48/177 [27%]; RR=1.50, 95% CI: 1.11–2.03; p=0.0077), and ≥10-point PSP improvement (63/176 [36%] vs 44/181 [24%]; RR=1.47, 95% CI: 1.06–2.04; p=0.018). Effects were consistent across symptoms, global-severity, and functional outcomes, with no evidence of heterogeneity between studies.

**Conclusions:** Across all three anchor criteria, roughly 1.5 to 1.75 times as many patients treated with roluperidone as with placebo reached thresholds of improvement that clinicians and patients would recognize as clinically meaningful, supporting its relevance for persistent negative symptoms in schizophrenia.

## 1. Introduction

A statistically significant (p<0.05) advantage over placebo on a psychometric scale in a randomized clinical trial (RCT) is reassuring because it indicates that the finding is unlikely to be a chance result: a 2-sided p value of 0.05 means that such data would arise only 5% of the time under the null hypothesis. Statistical significance, however, has only limited bearing on the clinical meaningfulness of the intervention. Regulators and health insurers who must make decisions about clinical meaningfulness and risk/benefit for groups of patients affected by a predefined condition typically rely on Cohen’s effect size (ES) and number needed to treat (NNT). For the individual patient and the prescribing clinician, the decision of whether a therapeutic intervention produces clinically meaningful improvement is a core element of their interaction and shared therapeutic decision. To justify a clinician’s recommendation and patient’s acceptance of a treatment, its effect must be large enough to alter the patient’s inner experience and be readily captured by the clinician’s general impression. The Clinical Global Impression of Severity (CGI-S) scale was designed to capture the clinician’s integrated judgment of overall illness severity incorporating symptoms, behavior, distress, and functioning into a single global assessment (Busner & Targum, 2007).

A 1-point improvement on the CGI-S is an empirically supported anchor for clinically meaningful improvement in schizophrenia. The criterion was derived by linking changes on symptom scales to clinician-rated global severity. Using equipercentile linking in a pooled database of 4,091 patients from seven pivotal antipsychotic trials, Leucht et al. (2005) found that a CGI-I rating of “minimally improved” corresponded to mean PANSS total score reductions of approximately 20% or more, depending on study duration. A complementary analysis supported the CGI-S threshold: a standardized mean difference of 0.46 corresponded to a 1-point between-group difference in CGI-S improvement, and CGI-S change was moderately correlated with standardized treatment effects (Pearson r=0.566; p<0.001; 95% CI: 0.431–0.676) (Damiani et al., 2026). Thus, a 1-point greater reduction in CGI-S severity with active treatment than control can be interpreted as clinically meaningful; if significantly more patients receiving active treatment than placebo reach this threshold, the treatment effect can be considered clinically meaningful.

A 7-to 10-point improvement on the PSP (range 0-100) is an empirically supported anchor for clinically meaningful functional improvement in schizophrenia. This range was derived from studies linking PSP change to clinically interpretable outcomes. Nasrallah et al. (2008), examining the measurement properties of the PSP total score in stable schizophrenia patients, estimated that an improvement of approximately 7 points represents detectable, clinically meaningful change. Nicholl et al. (2010), analyzing a relapse-prevention schizophrenia trial, found that a decline of 10 points or more on the PSP was strongly associated with clinical relapse, with 61% of relapses preceded by a decrement of this magnitude. Applied to treatment trials, where improvement rather than worsening is the relevant direction of change, these findings support a 7-to 10-point PSP gain as a clinically meaningful functional improvement. The present analysis uses the more conservative 10-point threshold.

Together, the CGI-S and PSP criteria provide complementary perspectives on clinical meaningfulness: the CGI-S criterion captures clinician-perceived global improvement, while the PSP criterion captures functionally meaningful change in domains relevant to daily functioning. Data obtained during a drug’s development program can help the prescriber judge whether its effect is large enough to benefit the individual patient.

The present manuscript applies the empirical criteria above to pooled responder analyses of the two registrational studies of roluperidone, a selective 5-HT2A/sigma-2/alpha-1A receptor antagonist targeting negative symptoms of schizophrenia. The studies are referred to here as Study 1 (Davidson et al., 2017) and Study 2 (Davidson et al., 2022); detailed descriptions of the trials can be found in those reports. Both studies were conducted as monotherapy, rather than as add-on to antipsychotics (Davidson et al., 2017, 2022), a design consistent with consensus recommendations for negative-symptom trials (Marder et al., 2020).

## 2. Methods

### 2.1. Response criteria and analysis sample

To assess the clinical meaningfulness of roluperidone’s treatment effect, we conducted responder analyses using data from the two trials. We applied three thresholds: a ≥20% improvement on the PANSS negative symptom factor score (NSFS), a ≥1-point CGI-S improvement (Damiani et al., 2026), and a ≥10-point PSP total-score improvement (Nasrallah et al., 2008; Nicholl et al., 2010). These thresholds were used to estimate the proportion of patients who achieved clinically meaningful improvement from baseline to Week 12. Analyses included only patients with a valid Week 12 assessment of the relevant endpoint. Responder status therefore reflected observed data rather than imputed or carried-forward values, and denominators differed slightly across endpoints. Pooling the two studies increased the precision of the responder estimates compared with either trial alone.

### 2.2. Statistical analysis

Responder proportions for roluperidone versus placebo were compared within each study and in a pooled analysis. Study-stratified (Mantel–Haenszel) estimates and tests of between-study heterogeneity are reported alongside the pooled results. Relative risks (RR) with 95% confidence intervals were calculated from the exact 2x2 responder counts, with confidence intervals derived on the log scale. The NNT was calculated as the reciprocal of the absolute risk difference.

## 3. Results

### 3.1. NSFS, CGI-S and PSP responder analyses

Responder rates on all three criteria were higher with roluperidone than with placebo, in each trial individually and in the pooled sample; the full set of counts, absolute differences, chi-square tests, relative risks and NNTs are presented in Table 1. In the pooled 12-week completers, roluperidone was superior to placebo on the ≥20% NSFS improvement criterion (RR=1.75, 95% CI: 1.25–2.45; p=0.0008; NNT 6.2), the ≥1-point CGI-S improvement criterion (RR=1.50, 95% CI: 1.11–2.03; p=0.0077; NNT 7.4) and the ≥10-point PSP improvement criterion (RR=1.47, 95% CI: 1.06–2.04; p=0.018; NNT 8.7). In relative terms, roluperidone-treated patients were therefore about 1.5 to 1.75 times as likely as placebo-treated patients to reach each threshold.

**Table 1.** Response at week 12 by study and for the two studies pooled (12-week completers).

| | Placebo | Roluperidone (%) | $\Delta$ (pts) | $\chi^2$ (p) | RR (95% CI) | NNT |
| --- | --- | --- | --- | --- | --- | --- |
| **Study 1 (placebo n=79, roluperidone n=79) 12-week completers** |  |  |  |  |  |  |
| $\geq 20\%$ NSFS decline | 9/54 (17%) | 18/54 (33%) | 17 | 3.88 (0.049) | 2.00 (0.99–4.05) | 6 |
| CGI-S 1-point decline | 13/54 (24%) | 22/54 (41%) | 17 | 3.43 (0.064) | 1.69 (0.95–3.00) | 6 |
| PSP 10-point improvement | 16/53 (30%) | 26/54 (48%) | 18 | 3.48 (0.06) | 1.59 (0.97–2.61) | 6 |
| **Study 2 (placebo n=172, roluperidone n=171) 12-week completers** |  |  |  |  |  |  |
| $\geq 20\%$ NSFS decline | 30/128 (23%) | 48/122 (39%) | 16 | 7.02 (0.008) | 1.68 (1.14–2.46) | 7 |
| CGI-S 1-point decline | 35/123 (28%) | 46/113 (41%) | 12 | 3.97 (0.046) | 1.43 (1.00–2.05) | 9 |
| PSP 7-point improvement | 37/128 (29%) | 50/122 (41%) | 12 | 4.58 (0.032) | 1.42 (1.00–2.00) | 9 |
| PSP 10-point improvement | 28/128 (22%) | 37/122 (30%) | 9 | 2.84 (0.092) | 1.39 (0.91–2.12) | 12 |
| **Studies 1 and 2 pooled** |  |  |  |  |  |  |
| $\geq 20\%$ NSFS decline | 39/182 (21%) | 66/176 (38%) | 16.1 | 11.15 (0.0008) | 1.75 (1.25–2.45) | 7 |
| CGI-S 1-point decline | 48/177 (27%) | 68/167 (41%) | 13.6 | 7.11 (0.0077) | 1.50 (1.11–2.03) | 8 |
| PSP 10-point improvement | 44/181 (24%) | 63/176 (36%) | 11.5 | 5.61 (0.018) | 1.47 (1.06–2.04) | 9 |

Notes: PSP 7-point change was unavailable for Study 1 because scores were recorded in deciles and therefore was not pooled. Pooled rows combine 12-week completers from Studies 1 and 2. NNTs are rounded up to the next whole number; exact pooled NNTs (6.2, 7.4 and 8.7) are given in the text. RRs and 95% CIs are based on exact 2×2 counts (CIs on the log scale), and NNTs on absolute risk differences. The $\chi^2$ column reports uncorrected chi-square statistics and p-values. Study-stratified Mantel–Haenszel pooled estimates were essentially unchanged (NSFS 1.75, 95% CI 1.25–2.46; CGI-S 1.50, 95% CI 1.11–2.04; PSP 1.46, 95% CI 1.06–2.02). Between- study heterogeneity was non-significant for all endpoints ( $I^2=0\%$ ), supporting pooling.

Absolute differences ranged from 11.5 to 16.1 percentage points. The largest absolute difference and the smallest p value were observed for the NSFS.

In clinical terms, approximately 2 in 5 patients treated with roluperidone achieved a ≥20% improvement in negative symptoms on the NSFS, approximately 2 in 5 achieved a clinically perceptible improvement in overall illness severity on the CGI-S, and approximately 1 in 3 achieved a 10-point functional improvement on the PSP, compared with approximately 1 in 4 patients on placebo on each criterion (Table 1). The effect was consistent across the two trials: study-specific relative risks ranged from 1.39 to 2.00, and no test of between-study heterogeneity approached significance (Table 1).

## 4. Discussion

Interpreting psychometric trial results through clinically anchored thresholds is useful for both drug development and clinical practice (Busner & Targum, 2007). For roluperidone, clinical meaningfulness is supported by the convergence of improvements across three independent anchors: PANSS negative symptoms, clinician-rated global severity, and functioning. The combined responder analyses presented here showed significantly higher rates of clinically meaningful improvement with roluperidone than placebo on all three endpoints—NSFS, CGI-S, and PSP. Because these thresholds were developed independently and assess different aspects of illness, their consistent results after 12 weeks strengthen the evidence that roluperidone’s effects are clinically meaningful.

To place the magnitude of roluperidone’s effect in context, Table 2 compares the relative risks and NNTs reported here with published benchmarks for approved antipsychotic and antidepressant drugs. In the pooled analysis, roluperidone showed relative risks of 1.75 for NSFS response, 1.50 for CGI-S response, and 1.47 for PSP response, with corresponding NNTs of 7, 8, and 9. These effects fall within the range reported for individual approved treatments. For antipsychotics, the benchmark relative risk versus placebo is 1.67 and the NNT is 5, although individual-drug effects vary substantially, with the least efficacious individual agents in that analysis, such as brexpiprazole, having an NNT of approximately 10 (Leucht et al., 2022). For antidepressants in major depressive disorder, the benchmark relative risk is approximately 1.35 (derived from the published odds ratio; see Table 2) and the NNT is 8, with individual drugs spanning a range that includes less favorable estimates such as reboxetine (relative risk approximately 1.20) (Cipriani et al., 2018). Roluperidone’s effects across NSFS, CGI-S, and PSP are therefore broadly comparable in magnitude to effects observed with approved psychiatric medications, although these comparisons are indirect and should be interpreted cautiously.

**Table 2.** Relative risks and numbers needed to treat: roluperidone compared with published benchmarks for approved treatments.

| Clinical response | RR vs placebo | NNT |
| --- | --- | --- |
| Roluperidone NSFS | 1.75 (1.25–2.45) | 7 |
| Roluperidone CGI-S | 1.50 (1.11–2.03) | 8 |
| Roluperidone PSP | 1.47 (1.06–2.04) | 9 |
| Antipsychotics <sup>(a)</sup> | 1.67 (1.59–1.73) | 5 |
| Antidepressants <sup>(b)</sup> | 1.35 (1.31–1.41)<sup>c</sup> | 8 |

<sup>a</sup> Leucht et al., 2022. <sup>b</sup> Cipriani et al., 2018. RR, relative risk. The antipsychotic RR is as published. An RR was not reported for antidepressants; the value shown (c) was derived from the published odds ratio of 1.70 assuming a placebo response rate of 37% (Furukawa et al., 2016) and is approximate.

## CRediT authorship contribution statement

All authors contributed to the conceptualization and design. JR conducted the data analysis. MD and JR drafted the manuscript, which was critically revised by SL and RL. All authors approved the final version.

## Acknowledgements

None.

## Data availability

Participant level data are not yet currently made available.

